# Atrial fibrillation, venous thromboembolism, and risk of pulmonary hypertension: A Swedish Nationwide Register Study

**DOI:** 10.1101/2024.06.25.24309502

**Authors:** Clara Hjalmarsson, Martin Lindgren, Niklas Bergh, Björn Hornestam, J. Gustav Smith, Martin Adiels, Annika Rosengren

**Affiliations:** Department of Cardiology, Sahlgrenska University Hospital, Gothenburg, Sweden; Department of Molecular and Clinical Medicine, Institute of Medicine at Sahlgrenska Academy, University of Gothenburg, Gothenburg, Sweden; Department of Medicine, Geriatrics and Emergency Medicine, Sahlgrenska University Hospital/Östra, Gothenburg, Sweden; School of Public Health and Community Medicine, Health Metrics Unit, University of Gothenburg, Gothenburg, Sweden; Department of Cardiology, Clinical Sciences, Lund University and Skåne University Hospital, Lund, Sweden; Wallenberg Center for Molecular Medicine and Lund University Diabetes Center, Lund University, Lund, Sweden

**Author notes:** **Address for correspondence** Clara Hjalmarsson, MD, PhD, Associate Professor of Cardiology, Dept. of Cardiology, Sahlgrenska University Hospital, University of Gothenburg, SE- 413 45, Gothenburg, Sweden. Deceased (Björn Hornestam).

**Keywords:** atrial fibrillation, pulmonary hypertension, venous thromboembolism

## Abstract

**Background:** Atrial fibrillation (AF) is a recognized risk factor for systemic arterial- and venous thromboembolism (VTE), including pulmonary embolism (PE), and may thereby contribute to the development of chronic thrombo-embolic pulmonary hypertension (PH). AF may also play a direct role in the development of post-capillary PH. We aimed to investigate the association between AF - with or without incident VTE - and the occurrence of PH.

**Methods:** A total of 521 988 patients diagnosed with AF between 1987–2013, without a previous diagnosis of VTE or PH, were identified from the Swedish National Patient Register (NPR) and matched for age, sex, and county with 1 017 277 population controls without AF, VTE or PH.

**Results:** The mean age of the AF patients was 71.1 (SD ±10.1) years and 42.8 % were women. During a median follow-up period of 11 (IQR 5.1,17) years, 4 454 (0.9%) AF patients and 1 855 (0.2%) controls were diagnosed with PH, HR 4.7 (4.4-5.0). The AF group had a significantly higher comorbidity burden at baseline, with a mean CHA_2_DS_2_-VASc of 2.9 compared to 2.1 in controls. In the absence of intercurrent VTE, the HR of PH was 4.2 (3.9-4.5) among AF patients compared to controls. Intercurrent VTE increased the HR of PH a further 1.9-fold (1.7-2.1) and 3.5 (3.1-4.0), among AF patients and controls, respectively. The HR for PH in AF patients with incident VTE was 8.1 (7.3-9.1).

**Conclusion:** AF was associated with a markedly increased risk of developing incident PH, and this risk was further increased by incident VTE.

## Introduction

Atrial fibrillation (AF) has been shown to induce a prothrombotic state and to promote inflammatory activity, thereby increasing the risk for both arterial and venous thromboembolism (VTE), including deep venous thrombosis (DVT) and pulmonary embolism (PE) ^1–4^.

Standard treatment of AF consists of drugs for rate or rhythm control, combined with oral anticoagulant medications (OAC) including DOAC (direct-acting oral anticoagulants) or warfarin ^4^. Several algorithms, such as the CHA_2_DS_2_-VASc score, are used to assess the clinical risk for ischemic stroke ^5^.

We have recently published data showing that the risk for VTE, and especially PE, is as high as for ischemic stroke during the first months after AF diagnosis, most markedly in younger patients and in women ^6^. Speculatively, the increased risk of VTE in AF may be mediated by multiple mechanisms, in accordance with Virchow’s triad^3^, including stasis in the caval and lung circulation with secondary in situ thrombosis, as well as dislodged thrombus from the right side of the heart. An association between AF and VTE has also been found in other studies ^7–9^. It is reasonable to assume that AF may enhance the risk of chronic thromboembolic pulmonary hypertension (CTEPH) over time, by multiple underdiagnosed subclinical pulmonary thromboembolic events.

Suboptimal resolution of peripheral thrombus, where endothelium interaction with thrombus may be phenotypically altered, and where genetic susceptibility may lead to modified fibrin structure and fibrinolytic resistance ^10, 11^ might play an essential role in the genesis of chronic pulmonary embolism and secondary pulmonary hypertension (PH) ^12–16^. The cumulative incidence of PH after a first episode of PE is estimated to be around 1% after 6 months, 3.1% after 1 year, and 3.8% after 2 years ^15^.

Apart from increasing the risk of CTEPH, AF can also lead to increased filling pressures which, if longstanding, may induce structural myocardial remodeling and atrial matrix reconstruction ^17^. This is *per se* a driving factor for atrial fibrosis, ventricular dysfunction, pulmonary vascular disease, and eventually secondary PH ^18^.

Thus, it is conceivable that AF could contribute to the onset of PH not only through elevated pre-capillary pulmonary pressures associated with subclinical pulmonary embolism and potential chronic thromboembolic pulmonary hypertension (CTEPH), but also by raising post-capillary pulmonary pressures. If left untreated, PH results in progressive right ventricular failure and ultimately death. Prognosis in PH is ominous, and especially with concomitant AF^19, 20^.

The primary aim of this study was to assess the occurrence of incident PH in patients with a diagnosis of AF compared to a matched control population without AF; a secondary aim was to investigate the association of incident VTE to PH in AF patients versus controls.

## Methods

### Study cohort

The study cohort was derived from the Swedish National Patient Register (NPR) which records all primary and secondary inpatient diagnoses in Sweden. The register has been in operation since 1960 and has full national coverage since 1987. In addition, since 2001, all outpatient visits to all hospitals in the country are also recorded in this NPR. Sweden provides universal healthcare to all citizens at a low cost which results in complete coverage of the Swedish population in NPR. Hospital diagnoses of major cardiovascular conditions in Sweden have been shown to have high validity (25).

From the NPR, we retrieved data on all first cases of AF as a primary or as a secondary diagnosis in any position identified in the NPR either through a discharge hospital code or as an outpatient code between January 1, 1987, and December 31, 2013, were retrieved. A first case was defined as no prior diagnosis of AF during the last 7 years. AF was defined as ICD codes 427D, and I48 for ICD9, and ICD10, respectively. All cases were matched by sex, age, and county to two control individuals without AF from the Swedish Population Register.

Data on several baseline comorbidities were also retrieved from the NPR and a comorbid condition was defined as present at baseline if there was at least one primary or secondary diagnosis in the 7 years preceding the AF diagnosis. ICD codes for baseline comorbidities are listed in supplementary **Table S1**. The Cause of Death register was used to record dates and causes of death.

AF cases were excluded together with their matched controls if vital data were inconsistent or if death occurred during the hospital stay. Controls with inconsistent vital data were removed. Control individuals were assigned the same inclusion date as their case.

The final data set was obtained by excluding individuals with prior (up to the same day as AF diagnosis) diagnosis of VTE and PH. See flow chart (supplementary **Figure 1**) for details on selection criteria for the study population.

### The National Prescribed Drug Register

Since mid-2005 all drug prescriptions in Sweden are recorded in the Prescribed Drug Register. For the cohort of AF patients and controls with inclusion dates between 2006-01-01 and 2013-12-31, data were retrieved from the prescribed drug registry on filled prescriptions of platelet inhibitors (Aspirin, Clopidogrel, Prasugrel), warfarin, and direct-acting oral anticoagulants (DOAC), as well as, other relevant drugs (ACE-inhibitors/angiotensin receptor blockers (ARB), Class I and III antiarrhythmic drugs, β-blockers, digitalis, diuretics, lipid lowering drugs, and COPD medications). Patients and controls were defined to be on treatment at baseline if there were at least two filled prescriptions for the drug within one year before the AF diagnosis. See supplementary **Table S2** for ATC codes.

### Definitions of outcomes

Pulmonary arterial hypertension (primary PH) was defined as a main diagnosis of 416A (ICD9) or I27.0 (ICD10). Secondary PH was defined as any diagnosis of 416C, 416W, 416X (ICD9) or I27.2, I27.8, I27.9 (ICD10).

### Statistical methods

Atrial fibrillation patients and matched control subjects were followed until i) a first event of PH, ii) death or, iii) end of study (2013-12-31). Events of VTE were recorded for time updated models.

Crude incidence rates and 95% confidence intervals (CI) were calculated using Poisson regression. Cox regression was used to assess the risk for PH following AF diagnosis. Unadjusted (only adjusted for age and sex) and adjusted (age, sex and baseline comorbidities: diabetes, hypertension, ischemic heart disease and heart failure) models were tested. Effect of incident VTE was included in the model as a time update when a VTE event occurred. To assess the interaction between AF and comorbidities, and the risk of PH we included interactions between AF and the comorbidities and used contrasts to compute specific hazard ratios.

In a separate cohort from 2006-01-01 to 2013-12-31, the models were further adjusted for the use of warfarin/DOAC **(shown in Table S3A and 3B),** aspirin or clopidogrel at baseline. The study was approved by the Regional Ethics Review Board in Gothenburg (Dnr. 104-15) and because the results are based on anonymized data the need for patient consent was waived. MA had full access to all the data in the study and takes responsibility for its integrity and the data analysis.

## Results

### Baseline

A total of 521,988 first AF cases (age 35-85) without a previous PH or VTE diagnosis were identified and matched for age and sex with 1,017,277 non-AF controls; the mean age was 71.1 (SD ± 10.1) and 71.0 (SD ±10.1) years for AF and controls, respectively. Heart failure (HF), chronic obstructive pulmonary disease (COPD), ischemic heart disease (IHD), adult congenital heart disease (ACHD), previous myocardial infarction (MI), valvular heart disease (VOC), and most other cardiopulmonary diagnoses and risk factors were more prevalent at baseline in the AF group, as was cancer (**Table 1)**. The AF population had higher CHA_2_DS_2_- VASc score compared to the control group, 2.9 compared to 2.1 respectively.

**Table 1.** Baseline characteristics in patients with atrial fibrillation and age- and sex-matched controls from the general population.

|  | **Atrial fibrillation** | **Population controls** |
| --- | --- | --- |
|  | n = 521 988 | n = 1 017 277 |
| Women | 223 337 (42.8) | 436 181 (42.9) |
| Age (years), mean (SD) | 71.1 (10.1) | 71.0 (10.1) |
| Heart failure | 119 889 (23.0) | 34 251 (3.4) |
| Chronic obstructive pulmonary disease | 30 886 (5.9) | 25 537 (2.5) |
| Valvular disease | 38 768 (7.4) | 11 690 (1.2) |
| Prior myocardial infarction | 60 046 (11.5) | 40 251 (3.9) |
| Ischemic heart disease | 135 577 (26.0) | 96 231 (9.5) |
| Congenital heart disease | 2 471 (0.5) | 873 (0.1) |
| Ischemic stroke | 57 610 (11.0) | 40 473 (4.0) |
| Transitoric Ischemic Attack | 21 100 (4.0) | 18 396 (1.8) |
| Haemorrhagic stroke | 4 582 (0.9) | 6 485 (0.6) |
| Hypertension | 158 112 (30.3) | 110 241 (10.8) |
| Diabetes | 70 482 (13.5) | 65 068 (6.4) |
| Renal failure | 14 790 (2.8) | 9 055 (0.9) |
| Asthma | 18 443 (3.5) | 16 889 (1.7) |
| Peripheral artery disease | 4 367 (0.8) | 3 942 (0.4) |
| Obesity | 7 711 (1.5) | 4 394 (0.4) |
| Cancer at baseline | 61 670 (11.8) | 96 303 (9.5) |
| CHA<sub>2</sub>DS<sub>2</sub>VASc, mean (SD) | 2.9 (1.7) | 2.1 (1.4) |

*Baseline characteristics of the whole study population; data shown as n (%) if not otherwise specified. For definitions (ICD codes), see supplementary table S1.*

### PH during follow-up

During a median follow-up of 11 [IQR 5.1,17] years, 4,454 (0.9 %) PH events were recorded in the AF group compared to 1,855 (0.2%) in the control group. Of these, 308 (0.06 %) and 141 (0.01 %) were primary PH (pulmonary arterial, hypertension, PAH), while 4,146 (0.8 %) and 1,714 (0.2 %) consisted of secondary or unspecified PH in the AF and control cohorts, respectively (**Table 2**).

**Table 2.** Registered diagnoses of PH in patients with atrial fibrillation and age- and sex-matched controls from the general population.

|  | **Atrial fibrillation** | **Population controls** |
| --- | --- | --- |
| **Pulmonary hypertension** |  |  |
| Events, total | 4 454 (0.9) | 1 855 (0.2) |
| Event rate | 75.2 (73.0-77.4) | 16.0 (15.3-16.7) |
| Follow-up, median [IQR], years | 11 [5.1,17] | 11 [5.2,17] |
| **Pulmonary arterial hypertension (group 1)** |  |  |
| Number of events (%) | 308 (0.06) | 141 (0.01) |
| Event rate | 5.2 (4.6-5.8) | 1.2 (1.0-1.4) |
| Follow-up, median [IQR], years | 11 [5.1,17] | 11 [5.2,17] |
| **Pulmonary hypertension (group 2-5)** |  |  |
| Number of events (%) | 4 146 (0.8) | 1 714 (0.2) |
| Event rate per 100,000 | 70.0 (67.9-72.2) | 14.8 (14.1-15.5) |
| Follow-up, median [IQR], years | 11 [5.1,17] | 11 [5.2,17] |

*Crude events, event rates per 100k person years (95% CI), and median follow-up time (years)*

Plots of cumulative events showed a greater risk for PH among AF cases compared to controls, **Figure 1**. In a Cox proportional hazard regression analysis, the hazard ratio for PH in AF cases vs non-AF controls was 4.7 (4.4-5.0), as shown in **Figure 2** (model adjusted for age and sex).

**Figure 1.**
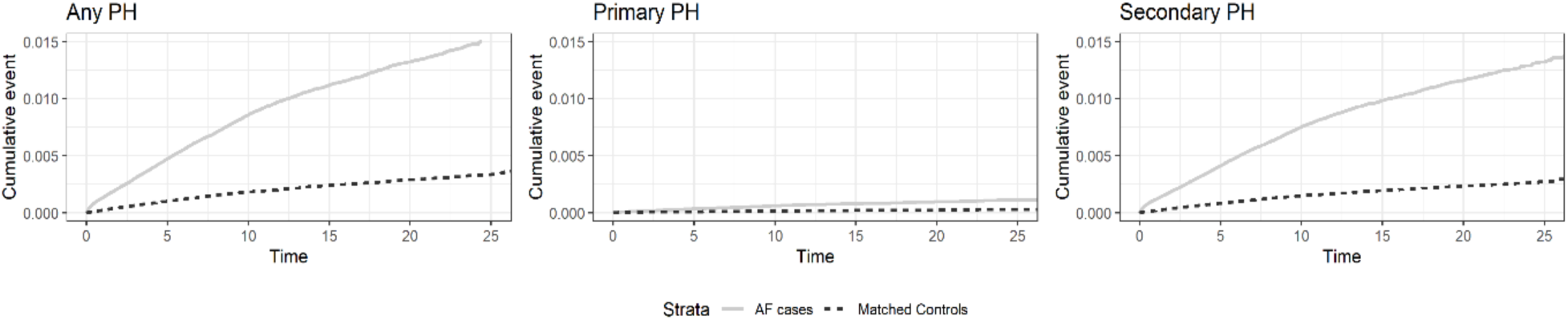
Plots of cumulative events PH in AF cases and matched controls. Y-axis represents cumulative events of PH without prior VTE.

**Figure 2.**
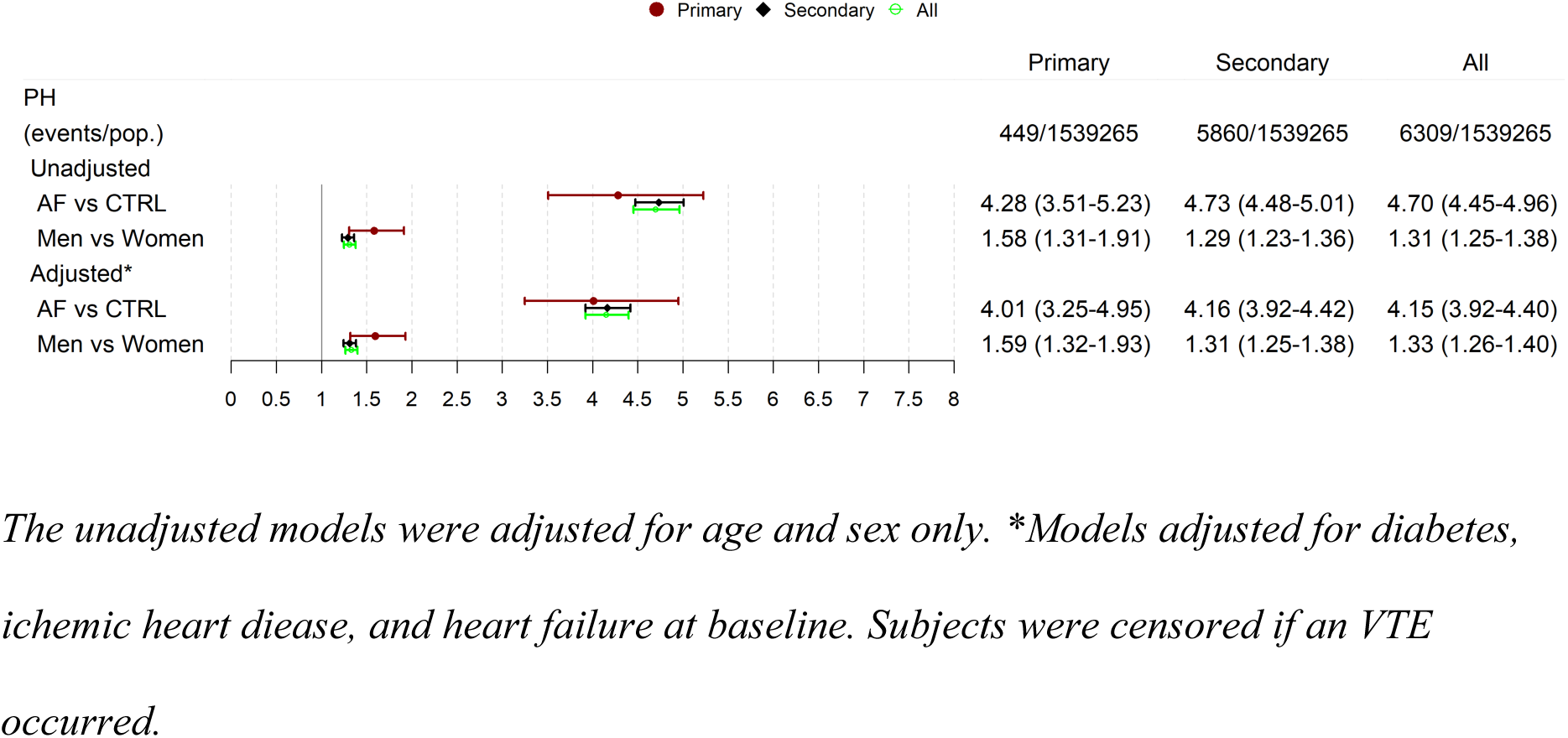
Hazard ratios for PH in AF cases and controls. The unadjusted models were adjusted for age and sex only. *Models adjusted for diabetes, ichemic heart diease, and heart failure at baseline. Subjects were censored if an VTE occurred.

### VTE as risk factor for PH

To elucidate the association of incident VTE to PH in AF patients and controls, we developed a time dependent Cox regression model. The model was updated when a subject had an incident VTE, thus allowing us to estimate the HR for PH with and without an incident VTE. There was a significant interaction between AF and incident VTE (*beta* −0.59, *p-value*< 0.001), see **Table 3**. Unadjusted results are shown in Supplementary **Table S4**. Using the model with an interaction between AF and VTE we could estimate the risk for PH in controls and AF patients. In controls with an incident VTE, the HR for PH was 3.5 (3.1-4.0), whereas AF patients experiencing an incident VTE had a HR of 1.9 (1.7-2.1) for developing PH. However, AF by itself was associated with a HR of 4.2 (3.9-4.5). Thus, relative to controls without incident VTE, AF patients with incident VTE had a HR of 8.1 (7.3-9.1), see **Table 3**. Unadjusted results are shown in Supplementary **Table S4**.

**Table 3.**
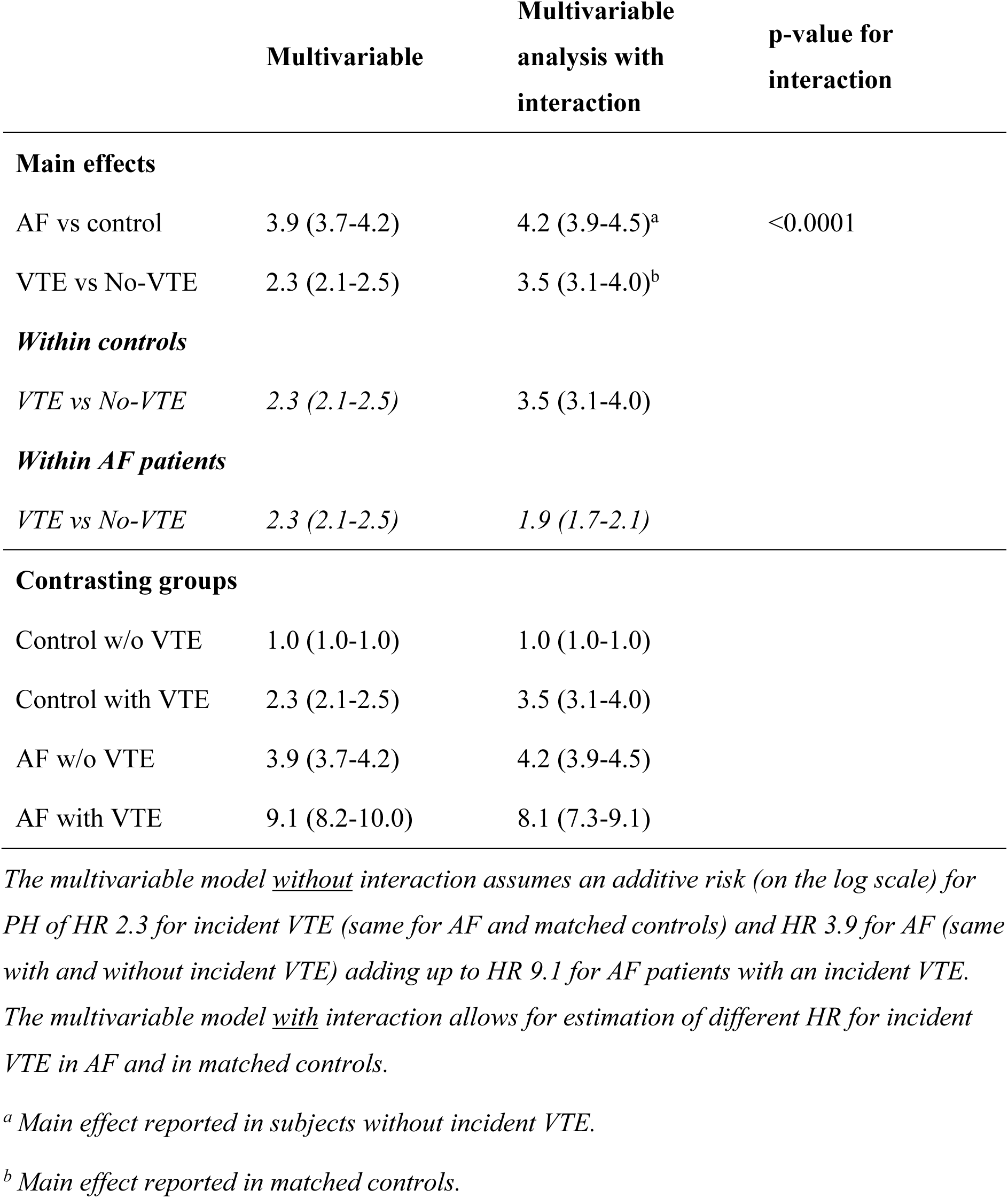
VTE as a risk factor for PH, adjusted for comorbidity (diabetes, hypertension, ischemic heart disease, and heart failure).

### Use of medication at baseline and follow-up

No data on drug prescriptions were available before the National Prescribed Drug register came into operation in mid-2005. Therefore, to assess the effect of medication at baseline, we investigated a subgroup diagnosed with a first episode of AF between 2006 and 2013, resulting in 197,177 AF patients and 383,012 controls. Baseline characteristics did not differ from the total study population, except for hypertension and cancer, which were more prevalent in both the AF and control groups (see **Table 4**). At baseline medication with OAC was more common in the AF group (see **Table S3A and S3B**).

**Table 4.** Baseline characteristics of subgroup with available data on baseline medication.

|  | **Atrial fibrillation** | **Population controls** |
| --- | --- | --- |
|  | n = 197 177 | n = 383 012 |
| Women | 81 068 (41.1) | 15 7951 (41.2) |
| Age, mean (SD) | 70.4 (10.3) | 70.3 (10.3) |
| Heart failure | 34 938 (17.7) | 10 429 (2.7) |
| Chronic obstructive pulmonary disease | 13 964 (7.1) | 11 500 (3.0) |
| Valvular disease | 16 768 (8.5) | 5 950 (1.6) |
| Prior myocardial infarction | 19 980 (10.1) | 13 576 (3.5) |
| Ischemic heart disease | 48 192 (24.4) | 37 651 (9.8) |
| Congenital heart disease | 1 236 (0.6) | 500 (0.1) |
| Ischemic stroke | 18 223 (9.2) | 13 214 (3.4) |
| TIA/Stroke | 8 126 (4.1) | 7 857 (2.0) |
| Haemorrhagic Stroke | 1 769 (0.9) | 2 366 (0.6) |
| Hypertension | 91 604 (46.5) | 69 704 (18.2) |
| Type 2 diabetes | 31 512 (16.0) | 31 416 (8.2) |
| Renal failure | 8 350 (4.2) | 4 946 (1.3) |
| Asthma | 7 941 (4.0) | 7 852 (2.0) |
| Peripheral artery disease | 1 805 (0.9) | 1 470 (0.4) |
| Obesity | 5 545 (2.8) | 3 244 (0.8) |
| Cancer at baseline | 32 037 (16.2) | 49 073 (12.8) |
| CHA2DS2VASc, mean (SD) | 2.87 (1.8) | 2.05 (1.5) |
| Treatment with anticoagulants at baseline | 24 641 (12.5) | 3 540 (0.9) |

*Data shown as n (%) if not otherwise specified.*

The pattern for PH in this subgroup was similar to the one observed in the total study population. Further adjusting the models for the use of medications at baseline provided marginally changed risk estimates for PH, compared to the total population; the most noticeable change was observed in the effect of incident VTE on PH (see **Table 5**).

**Table 5.** VTE as a risk factor for PH, adjusted for comorbidity (diabetes, hypertension, ischemic heart disease, and heart failure) and for treatment with anticoagulants*.

|  | **Multivariable** | **Multivariable**<br>**with interaction** | **Multivariable**<br>**with interaction,**<br>**adjusted for**<br>**treatment** |
| --- | --- | --- | --- |
| **Main effects** |  |  |  |
| AF vs CONTROL | 5.43 (4.87-6.05) | 5.82 (5.19-6.53)<sup>a</sup> | 4.49 (3.94-5.12)<sup>a</sup> |
| VTE vs No-VTE | 2.46 (2.08-2.91) | 4.20 (3.15-5.60)<sup>b</sup> | 4.12 (3.09-5.49)<sup>b</sup> |
| **Within controls** |  |  |  |
| VTE vs No-VTE | 2.46 (2.08-2.91) | 4.20 (3.15-5.60) | 4.12 (3.09-5.49) |
| **Within AF patients** |  |  |  |
| VTE vs No-VTE | 2.46 (2.08-2.91) | 1.97 (1.60-2.43) | 2.06 (1.66-2.54) |
| **Contrasting groups** |  |  |  |
| Control w/o VTE | 1.00 (1.00-1.00) | 1.00 (1.00-1.00) | 1.00 (1.00-1.00) |
| Control with VTE | 2.46 (2.08-2.91) | 4.20 (3.15-5.60) | 4.12 (3.09-5.49) |
| AF w/o VTE | 5.43 (4.87-6.05) | 5.82 (5.19-6.53) | 4.49 (3.94-5.12) |
| AF with VTE | 13.34 (10.94-16.28) | 11.48 (9.16-14.38) | 9.23 (7.32-11.64) |

\*See table 3 for full description

## Discussion

In this national Swedish register study of patients with AF, the most important findings were: 1) over time, AF was found to be associated with a 4.7-fold increase in the risk of developing PH independently of any known VTE events. This suggests that AF itself may play a role in the pathogenesis of PH; 2) a history of incident VTE in patients with AF further increased the risk of developing PH two-fold. This indicates that both post-capillary and pre-capillary mechanisms leading to PH may be involved.

Even when adjusting for age and sex, the comorbidity burden in the atrial fibrillation (AF) population was notably higher compared to controls. This reinforces the notion that AF may serve as an indicator of overall poor health. Consequently, given the more pronounced cardiovascular and pulmonary comorbidities, it is expected that patients with atrial fibrillation (AF) would have a higher risk of developing secondary PH. However, the risk for PH remained significantly elevated even after adjusting for relevant comorbidities, indicating that AF itself is associated with the occurrence of PH.

The hemodynamics of patients with AF is characterized by increased diastolic filling pressures, atrial enlargement, and fibrosis, as well as fluctuating diastolic filling time, systolic ventricular stroke volume, and ejection fraction (EF) from beat to beat. Furthermore, lack of synchrony between the atria and ventricles results in fluctuating pressures, volume, and blood flow through the caval and pulmonary circulation ^21^. Also, mitral- and/or tricuspid insufficiency of varying size is common in AF. AF *per se* also induces atrial and ventricular enlargement, which adds to the above. These complex hemodynamic changes could lead to increased pericardial restraint and ventricular interdependence, abnormal right ventricle – pulmonary artery coupling, and negative effects on pulmonary vasculature ^18^. AF may be both a cause and an effect of heart failure, irrespective of ejection fraction, but the exact contribution of different types of heart failure to the development of PH in this patient cohort was not possible to assess. However, we found that the combination of AF with either heart failure or valvular heart disease further increased the risk for PH approximately 8 to 10-fold.

The second main finding, namely that an incident diagnosis of VTE doubled the risk for PH in AF patients, is in line with the discussion above. It is crucial to highlight that this is in addition to the 4.7-fold increased risk associated with AF alone. Therefore, the combination of AF and VTE results in an almost 10-fold increased risk for developing PH.

AF induces a prothrombotic state by potentiating all three mechanisms of Virchow’s triad, as described in detail by Watson et al ^3^. Abnormal changes in blood flow (blood stasis), abnormalities in vessels and myocardial walls (endothelial dysfunction), as well as in blood constituents (hemostatic and platelet activation, inflammation and growth factor changes) increase the risk for thromboembolism ^2–4^, thereby contributing to VTE development ^6^. Furthermore, in a patient with AF, a pulmonary embolism may be misdiagnosed as a pulmonary infection, an asthma or COPD exacerbation, or heart failure, and therefore might go undetected. Less than half of the patients diagnosed with pulmonary embolism receive a systematic screening for deep vein thrombosis (DVT), while a significant portion of patients with DVT have an undetected PE ^22^. Thus, the prothrombotic effects of AF, combined with its detrimental impact on cardiac and pulmonary hemodynamics, may significantly increase the risk of PH occurrence.

We found a nearly 14-fold higher risk of PH in patients with both AF and chronic obstructive pulmonary disease (COPD). In COPD patients, PH typically results from parenchymal lung changes ^23–25^, but there is also a documented increased risk of thrombosis within the pulmonary vasculature due to inflammation ^26^. Furthermore, the combined impact of atrial fibrillation’s prothrombotic and hemodynamic effects ^27^ could potentially amplify the risk of PH development in these patients.

Contemporary management of AF includes early initiation of anticoagulation based on CHA_2_DS_2_-VASc score and other relevant prothrombotic risk factors. However, because this study encompassed a broad time frame, there was significant variability in treatment indications and therapy choices for AF across different periods. To tackle this issue and ensure a more homogeneous study population, a subgroup analysis was conducted specifically on patients treated with anticoagulants, yielding comparable findings.

Our findings highlight the need of better understanding the intricate relationship between AF and PH, especially in the context of other cardiopulmonary comorbidities, since the combination of these conditions generally portends a much worse prognosis. Timely treatment of AF by adequate rhythm and/or rate control, including catheter ablation, and anticoagulation when indicated, is considered essential in reducing the risk of thromboembolic complications and/or heart failure according to current recommendations. Our findings underscore that AF itself significantly raises the risk of developing incident PH, a widely recognized indicator of poor prognosis, reinforcing the importance of initiating prompt and aggressive therapy for AF.

### Strengths and limitations

Major strengths of the present study include a large number of AF cases and population-based controls, near-complete coverage of the Swedish population, and well-validated data on diagnoses, medication, outcome, and confounders. Some limitations are that: data were obtained from several databases and were not primarily collected for research purposes; the incidence of PH is likely to be underestimated to an unknown but probably substantial extent, since PH may be underdiagnosed and/or less often recorded in patient notes; the use of routine echocardiography was much less common during the initial time of our data collection than today, which might have contributed to underestimation of the PH diagnosis; additionally, some PH diagnoses might have been erroneously reported as PAH, since the incidence of PAH in our population is quite high in comparison to other published epidemiological data.

Furthermore, we lack data on adherence to OAC treatment which is a limitation since there is a potential link between poor adherence and risk for thromboembolism. Unknown confounders, such as silent/undiagnosed AF among controls, may also be present, but the interference risk would be low, given the very large sample size.

Additional points to consider include that our data were collected between January 1, 1987, and December 31, 2013. During this period, large changes in the treatment of AF based on validated risk scores (CHA_2_DS_2_ and CHA_2_DS_2_-VASc) were widely implemented, and mostly during the “pre-DOAC” era. It is highly likely that the initiation and prevalence of OAC treatment in these patients have significantly increased compared to previous periods.

## Conclusion

In this study, which included a large population of patients with AF and age- and sex-matched controls, we observed that AF alone is linked to a 4.7-fold higher risk of developing incident PH, in the absence of prior VTE. Additionally, the occurrence of new VTE events significantly elevates this risk to nearly 10-fold. Established causes of PH, such as heart failure, valvular heart disease, and COPD further increase the risk in AF patients. This association is probably mediated by several mechanisms where AF-related hemodynamic consequences and AF-induced prothrombotic effects may play important roles. Further research is needed to elucidate the mechanisms of AF-induced pulmonary vascular disease and to identify which AF patients are most susceptible to developing PH.

## Data Availability

Original data are available through contact with Dr. Martin Adiels, responsible for the statistical analysis of the study.

## Acknowledgements

This work was supported by grants from the following: the Swedish state under an agreement between the Swedish Government and the County Councils Concerning Economic Support of Research and Education of Doctors [grant number ALFGBG-427301, ALFGBG-966211, ALFGBG-971608, ALFGBG-979104]; the Swedish Heart and Lung Foundation [grant number 2015-0438, 2018-0589, 2021-0345]; the Swedish Research Council [2018-02527, VRREG 2019-00193]

---

**Supplementary figure 1.**
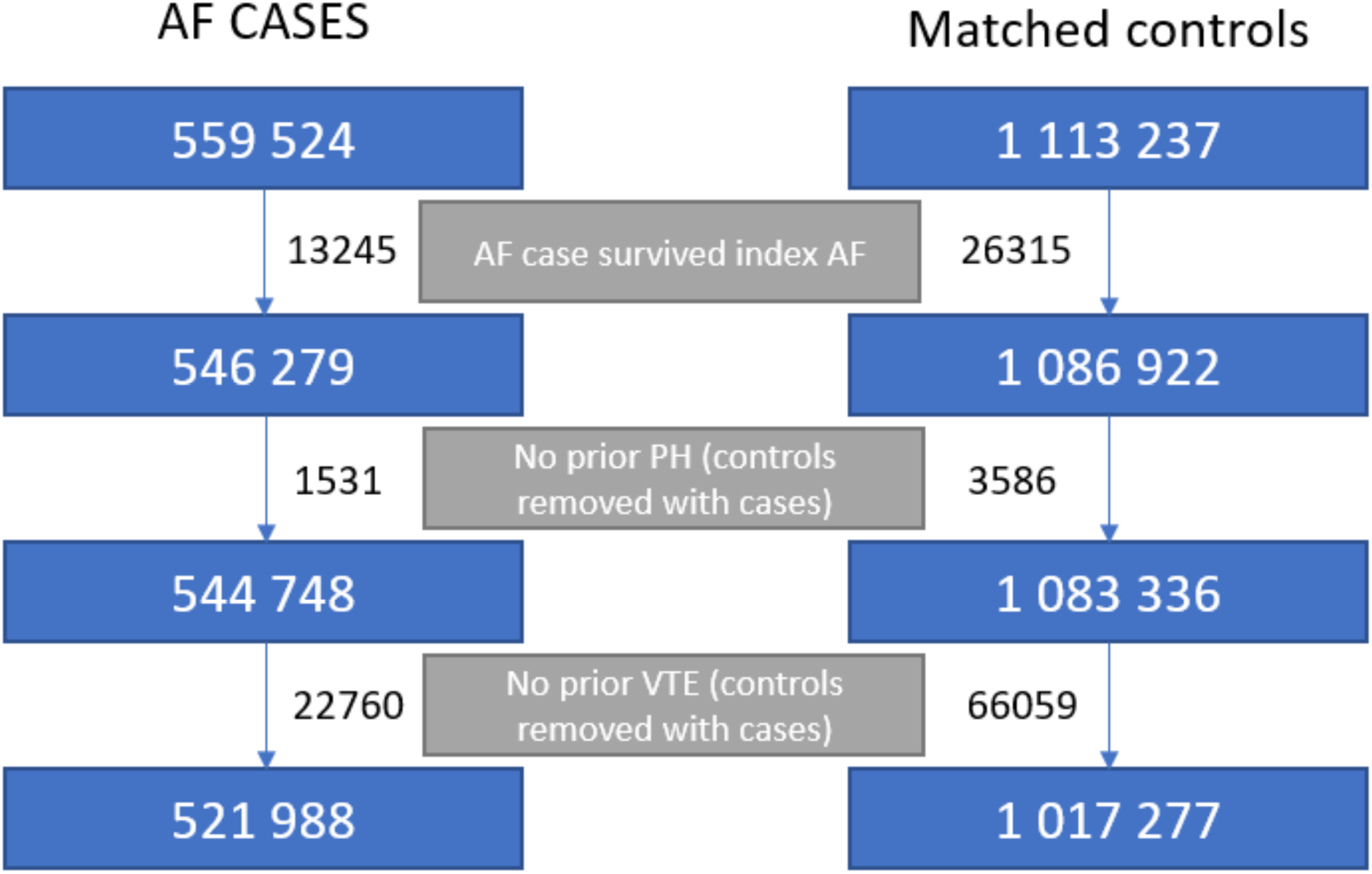
Flow chart showing selection of study population.

**Supplementary Table 1 (S1).** List of ICD-codes used in the study.

|  | ICD-9 | ICD-10 |
| --- | --- | --- |
| Hypertension | 401-405 | I10-I15 |
| Ischaemic heart disease | 410-414 | I20-I25 |
| Myocardial infarction | 410 | I21 |
| Valvular heart disease | 394-398, 424 | I05-I09, I34, I35 |
| Congestive heart failure | 428 | I50 |
| Pulmonary embolism | 415B | I26 |
| Congenital heart disease | 745-747, 758A, 758G, 759W | Q20-Q28, Q87, Q90, Q96 |
| Ischaemic stroke | 434, 436 | I63, I64 |
| TIA* | 435 | G45 |
| Cerebral hemorrhage | 431,432 | I61, I62 |
| Deep venous thrombosis | 451B, 453 | I80 |
| Peripheral Artery Disease | 443X | I839B |
| Diabetes mellitus | 250 | E10-E14 |
| Obesity | 278A, 278B | E65, E66 |
| Renal failure | 584-587 | N17-N19 |
| COPD<sup>#</sup> | 490-492 | J40-J44 |
| Asthma | 493 | J45 |
| Cancer | 140-209 | C00-C99 |

*\*Transitory ischaemic attack, <sup>#</sup>Chronic obstructive pulmonary disease*

**Supplementary Table 2 (S2).** List of ATC-codes used in the study.

| **Compound name** | **ATC code** |
| --- | --- |
| Warfarin | B01AA03 |
| Rivaroxaban | B01AF01 |
| Apixaban | B01AF02 |
| Pradaxa | B01AE07 |

**Supplementary Table 3A (S3A).** Prevalence of OAC at baseline and within 30 days, 90 days, and one year after diagnosis, data shown as % (n).

| **Time point** | **AF** | **CTRL** |
| --- | --- | --- |
| Baseline | 12.5% (24 641) | 0.9% (3 540) |
| Within 30 days | 47% (92 604) | 1% (3 796) |
| Within 90 days | 51.5% (101 524) | 1.1% (4 265) |
| One year | 55.6% (109 645) | 1.5% (5 764) |

**Supplementary Table 3B (S3B).**
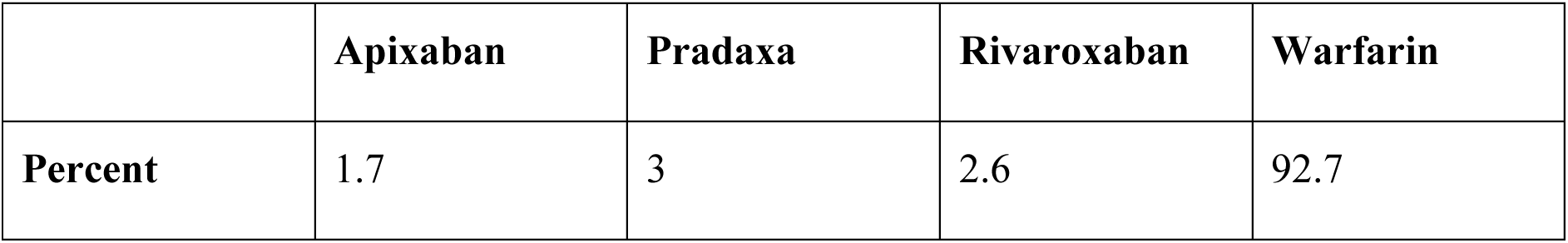
Type of OAC used by study population, data shown as %.

|  | **Apixaban** | **Pradaxa** | **Rivaroxaban** | **Warfarin** |
| --- | --- | --- | --- | --- |
| **Percent** | 1.7 | 3 | 2.6 | 92.7 |

**Supplementary Table 4 (S4).**
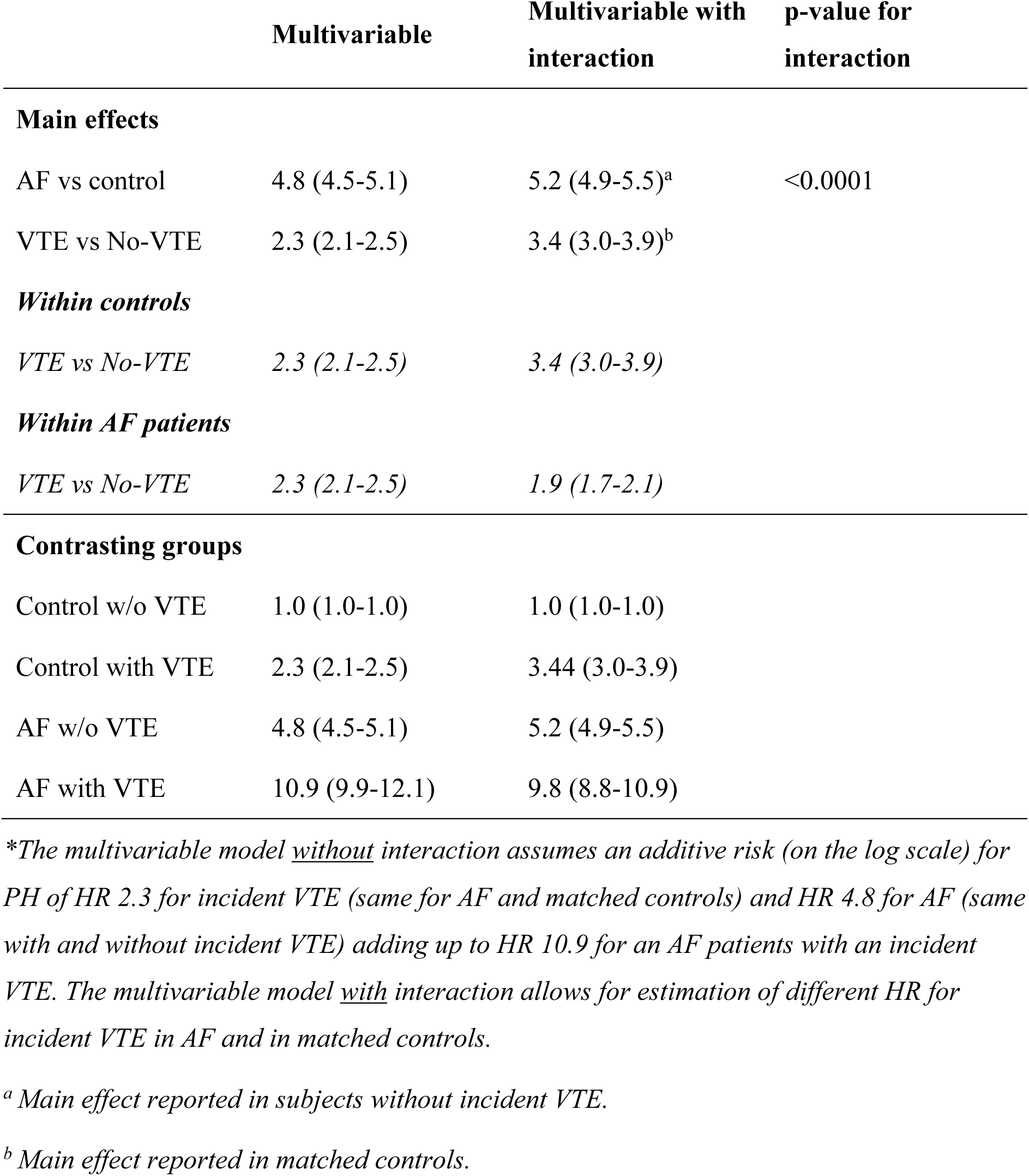
Hazard ratio for incident VTE as a risk factor for PH (age and sex adjusted)*.

|  | Multivariable | Multivariable with interaction | p-value for interaction |
| --- | --- | --- | --- |
| **Main effects** |  |  |  |
| AF vs control | 4.8 (4.5-5.1) | 5.2 (4.9-5.5)<sup>a</sup> | <0.0001 |
| VTE vs No-VTE | 2.3 (2.1-2.5) | 3.4 (3.0-3.9)<sup>b</sup> |  |
| **Within controls** |  |  |  |
| VTE vs No-VTE | 2.3 (2.1-2.5) | 3.4 (3.0-3.9) |  |
| **Within AF patients** |  |  |  |
| VTE vs No-VTE | 2.3 (2.1-2.5) | 1.9 (1.7-2.1) |  |
| **Contrasting groups** |  |  |  |
| Control w/o VTE | 1.0 (1.0-1.0) | 1.0 (1.0-1.0) |  |
| Control with VTE | 2.3 (2.1-2.5) | 3.44 (3.0-3.9) |  |
| AF w/o VTE | 4.8 (4.5-5.1) | 5.2 (4.9-5.5) |  |
| AF with VTE | 10.9 (9.9-12.1) | 9.8 (8.8-10.9) |  |

\*The multivariable model without interaction assumes an additive risk (on the log scale) for PH of HR 2.3 for incident VTE (same for AF and matched controls) and HR 4.8 for AF (same with and without incident VTE) adding up to HR 10.9 for an AF patients with an incident VTE. The multivariable model with interaction allows for estimation of different HR for incident VTE in AF and in matched controls.
<sup>a</sup> Main effect reported in subjects without incident VTE.
<sup>b</sup> Main effect reported in matched controls.

